# Transcriptional Signatures of the Right Ventricle in End-Stage Heart Failure

**DOI:** 10.1101/2024.10.22.24315927

**Authors:** Jonah D. Garry, Giovanni E. Davogustto, Vineet Agrawal, Fei Ye, Kelsey Tomasek, Yan Ru Su, Tarek Absi, James D. West, Anna Hemnes, Evan L. Brittain

**Affiliations:** Division of Cardiovascular Medicine, Vanderbilt University Medical Center, Nashville, TN; Department of Veteran Affairs, Tennessee Valley Healthcare System, Nashville, TN; Department of Biostatistics, Vanderbilt University Medical Center, Nashville, TN; Department of Cardiac Surgery, Vanderbilt University Medical Center, Nashville, TN; Division of Allergy, Pulmonary, and Critical Care Medicine, Vanderbilt University Medical Center, Nashville, TN

## Abstract

The molecular mechanisms of right ventricular (RV) adaptation to stress and failure in end stage heart failure (HF) are largely unknown. In this study, we performed transcriptomic analysis on paired RV and left ventricular (LV) myocardial samples from 33 subjects with AHA stage D HF referred for transplantation, and 8 controls who provided donor hearts not ultimately used for transplantation. We categorized patients by RV function - defining failure by Pulmonary Artery Pulsatility Index <1.85 - into control (n=8), compensated (n=25), and failing groupings (n=8). Our analysis found dysregulation of extracellular matrix remodeling, fatty acid metabolism, mitochondrial function, and inflammation in the stressed RV (comparing compensated and failing RVs to controls), implicating similar molecular mechanisms to those identified from studies of pre-capillary pulmonary hypertension (PH) associated RV dysfunction. The compensated and failing RVs were differentiated by protein production/processing and inflammation. PPAR signaling and fatty acid metabolism were consistently enriched in the RV compared to the LV. We also examined 16 previously proposed transcriptional biomarkers of RV failure, finding that none effectively differentiated between RV compensation and failure, and only 4 were RV-specific (changing in RV-failure but not LV-failure). Our findings suggest that the molecular mechanisms of pre-capillary PH and HF-associated RV dysfunction exhibit significant overlap. Furthermore, fatty acid metabolism and related signaling may be constitutively different between the RV and LV in both normal function and failure.

## Introduction

Right ventricular (RV) dysfunction is common in heart failure (HF) and consistently associated with adverse outcomes,^1–5^ yet the molecular mechanisms underlying RV compensation and progression to failure are incompletely understood.^6^ There are currently no RV-specific biomarkers or medical therapies. The pathways thus far identified associating with RV failure derive predominantly from animal models and human studies of pulmonary arterial hypertension (PAH) with changes noted in inflammation,^7–9^ sex hormone expression,^10,11^ cellular metabolism,^12,13^ and extracellular matrix (ECM) organization.^9,14^ Multiple outstanding questions remain, including how these pathways relate to progression from RV compensation to failure, whether pathways associated with RV failure in PAH are also associated with RV failure in patients with HF, and to what degrees the pathways identified are unique to the RV.

Myocardial transcriptomic analysis is a powerful tool for understanding molecular mechanisms of cardiac failure that has been deployed extensively to understand cellular changes in the failing left ventricle (LV) in HF.^15^ In contrast, few human studies have examined transcriptomic changes associated with RV progression from compensation to failure with a focus on biomarker development,^14,16^ one in PAH and one in HF. No studies have directly compared transcriptional changes in RV failure compared to LV failure.

To address these gaps, we utilized bulk RNA-sequencing of myocardial tissue to describe transcriptional changes that occur in the RV 1) In the context of stress from left-heart failure by comparison to controls and 2) As the RV progresses from compensation to failure. We further compared transcriptomic changes that occur in RV failure to the transcriptomic changes that occur in LV failure. We hypothesized that comparison of the failing RV to the failing LV would yield insights into the unique and overlapping molecular mechanisms of ventricular failure.

## Methods

All procedures were reviewed and approved by the Vanderbilt Institutional Review Board # 090828 (biorepository registry) and 181788. All patients provided written informed consent for participation in the biorepository registry with sample collection and subsequent molecular analysis.

Myocardial tissue samples were obtained from the Vanderbilt Heart and Vascular Institute Biorepository. Hearts were obtained from patients diagnosed with AHA Stage D HF who were undergoing orthotopic heart transplant without regard for left ventricular systolic function or underlying HF etiology. We excluded any patients that had a left ventricular assist device prior to transplant, a prior heart transplant, congenital heart disease, or a clinically suspected hereditary cardiomyopathy. Samples without paired LV and RV tissue available were excluded. Control samples were obtained from unmatched donor hearts that were not ultimately used for transplantation due to intravenous drug use, abnormal T waves during stroke, possible pericarditis, known coronary artery disease, and/or history of smoking. In all hearts, myocardial samples free of macroscopic fibrosis and epicardial fat were obtained from the RV and LV free-walls at 5-7cm above the apex, flash frozen in liquid nitrogen and stored in - 80 °C freezers. Control samples were dissected and immediately frozen in the operating room at the time of explant. For HF samples, the time from cross clamp to sample freezing (cold ischemic time) was available in 30 of the 33 subjects with HF, with a median time 47 minutes (IQR 38 – 60).

### RNA extraction and sequencing

Approximately 90 to 120 mg of tissue per ventricle per study subject was used for RNA extraction. Total RNA was isolated using the RNeasy miRNA Mini Kit and the QIAcube (Qiagen) for nucleic acid extraction following the manufacturer’s instructions. Total RNA was submitted to the Vanderbilt VANTAGE Core for RNA sequencing. Sample quality and concentration of the total RNA were determined using the 2100 Bioanalyzer, RNA ScreenTape®, and Tapestation Analysis Software A.02.02(SR1) (Agilent Technologies; Santa Clara, CA). For each library, approximately 100 ng of total RNA underwent ribosomal RNA depletion. Then, cDNA libraries were prepared using stranded mRNA (polyA-selected) with the TruSeq RNA Sample Prep Kit (Illumina; San Diego, CA) according to the manufacturer’s instructions. Library quality was assessed with the 2100 Bioanalyzer. Sequencing was performed in a single batch, at a Paired-End 150 bp on the Illumina NovaSeq6000. RNAseq data were quality controlled repeatedly at raw, alignment, and expression level.^17^ QC3 was used for quality control of raw and alignment data.^18^ Alignments were performed with Tophat 2 against the HG19 human reference genome.^19^

### Group Definitions and Comparisons

Baseline demographics were collected for all patients. In HF patients, the laboratory, echocardiographic, and hemodynamic parameters closest to time of transplant were recorded. We defined RV failure by a pulmonary artery pulsatility index (PAPi) of <1.85 based on clinical association with adverse outcomes in end-stage heart failure^20–22^ and evidence of RV myofilament contractile dysfunction at this cutoff.^23^ We categorized myocardial samples into 5 mutually exclusive groupings: 1) Control RV (n = 8); 2) Failing RV (n = 8); 3) Compensated RV (n=25); 4) Control LV (n=8); 5) Failing LV (n=33). Definitions for each of these groupings is displayed in **Table 1**.

**Table 1:** Key Definitions.

| **Table 1: Key Definitions** |  |
| --- | --- |
| **Pulmonary Artery Pressure Index (PAPi)** | $PAPi = (sPAP - dPAP) / RAP$ |
| **Failing RV** | RV sample from a patient who underwent transplantation for clinically established AHA Stage D heart failure<br>$PAPi < 1.85$ |
| **Compensated RV** | RV sample from a patient who underwent transplantation for clinically established AHA Stage D heart failure<br>$PAPi \geq 1.85$ |
| **Control RV** | RV sample from heart harvested from brain dead subject with initial plan for transplantation, however ultimately declined |
| **Failing LV** | LV sample from a patient who underwent transplantation for clinically established AHA Stage D heart failure |
| **Control LV** | LV sample from heart harvested from brain dead subject with initial plan for transplantation, however ultimately declined |
| **RV Under Stress** | RV in a patient with clinically established AHA Stage D heart failure (regardless of Failing or Compensated RV status) |
| sPAP = systolic pulmonary artery pressure; dPAP = diastolic pulmonary artery pressure; RAP = right atrial pressure; RV = Right Ventricle; LV = Left Ventricle |  |

We sought to understand myocardial transcriptional changes in the RV through four sets of comparisons. 1) We examined the RV under stress by comparing Control RV to Compensated RV and Control RV to Failing RV; 2) We described differences between the Compensated RV and Failing RV; 3) We identified transcriptional changes unique to the failing RV as compared to the failing LV by examining Failing RV vs Control RV compared to Failing LV vs Control LV. Of note, for these comparisons we only included Failing LV samples from patients with concurrent RV failure. 4) Lastly, we identified transcriptional differences between the RV and LV in Control samples and HF sample. We then compared the differences between ventricles in controls and heart failure. **Figure 1** displays the study processes and overview of transcriptional comparisons.

**Figure 1:**
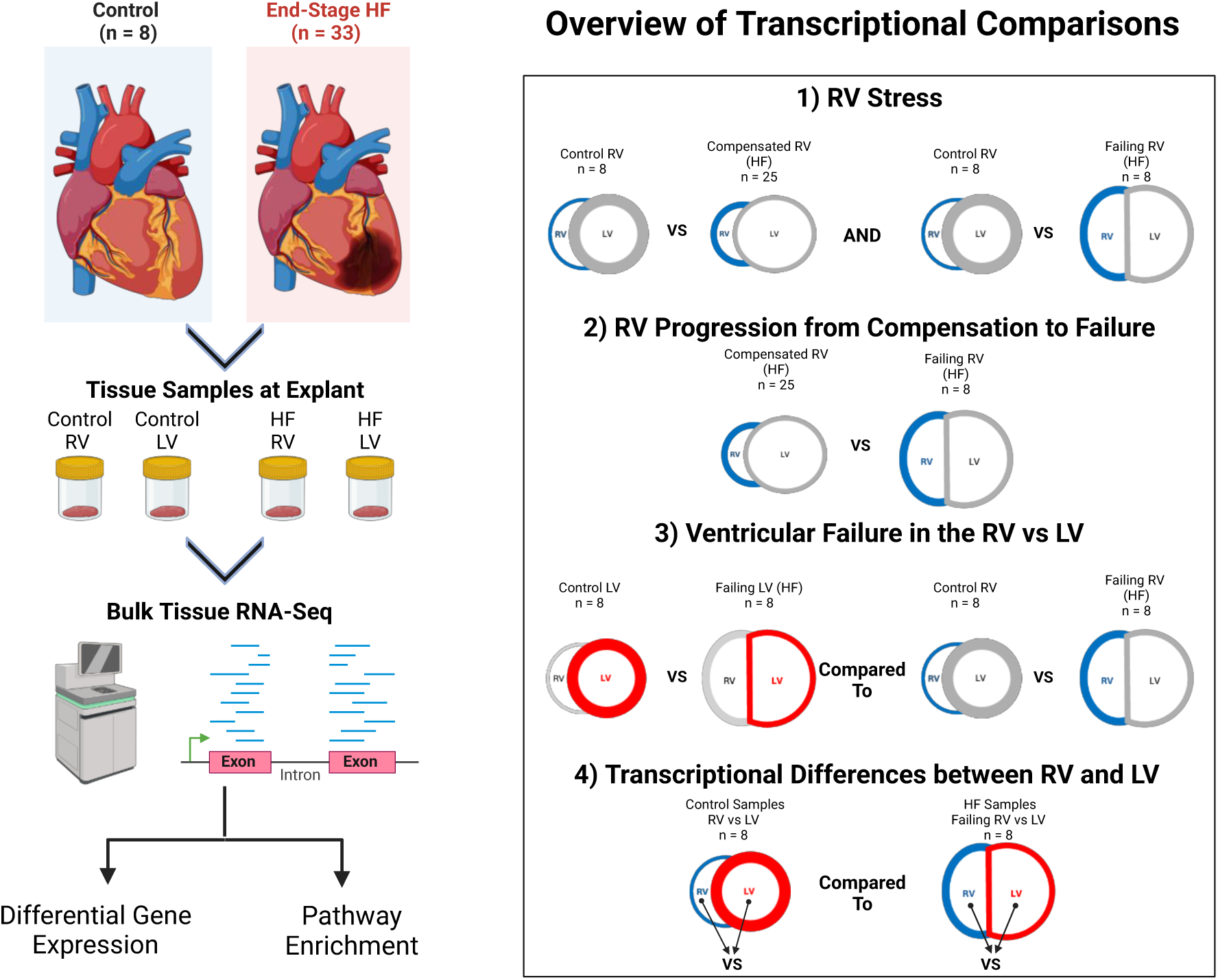
Study Procedures Figure 1 displays the overall study design, and an overview of the transcriptional comparisons performed. Figure created in BioRender.com.

### RV-Specific Biomarker Candidates

We examined previously proposed biomarkers of RV failure from two prior myocardial transcriptomic studies for their ability to discriminate between control, compensated, and failing RV samples. An optimal RV-specific biomarker in heart failure would change minimally between normal LV and failing LV, therefore we compared whether normalized gene counts changed in both the RV and LV (comparing control to failing ventricles). To identify novel potential biomarkers of RV failure, we described the top 10 genes that were differentially expressed between failing and control RV with statistical significance, but were not statistically significantly differentially expressed between failing and control LV.

### Statistical Analysis

Descriptive data are presented as count and frequencies for categorical variables and quartiles for continuous variables, unless otherwise specified. Raw count data was normalized and transformed to the log2 scale prior to differential expression analysis using the R package DESeq2.^24^ Dispersion estimates plots were visually inspected to ensure appropriate fit of the negative binomial model. To visualize separation of groups we performed unsupervised clustering analyses via principal components analysis (PCA) and supervised clustering by partial least squares discriminant analysis (PLS-DA) to maximize separation. We identified differentially expressed genes (DEGs) based on a Benjamin-Hochberg false discovery rate (FDR) adjusted p-value of <0.05 and an absolute log fold change of 0.5 of greater. We performed functional analysis using Overrepresentation Analysis (ORA) on gene sets of interest and compared between groups using Gene Set Enrichment Analysis (GSEA) on the WEB-based Gene Set AnaLysis Toolkit (WebGestalt) 2024.^25^ For ORA and GSEA, we selected the top 20 KEGG pathways and Gene Ontology Biological Processes with FDR-adjusted p-value <0.10, excluding KEGG pathways for non-cardiac disease-specific processes (i.e pathways for “Alzheimer Disease” or “Leishmaniasis. For biomarker assessment, we identified genes with significantly different normalized counts based on a Bonferroni-adjusted (for number of biomarkers assessed) p-value of <0.05. All statistical analyses were performed with R software program, version 4.3.3.

## Results

### Patient characteristics

We included 33 subjects with AHA stage D HF and 8 controls from the Vanderbilt Heart and Vascular Institute Biorepository. Subjects with HF were older at harvesting (52.7 years vs 43.5 years) with a higher proportion of males (64% vs 50%). Other than age, clinical characteristics are not available for control subjects. Laboratory, echocardiographic, and hemodynamic parameters closest to transplant for the HF group are displayed in **Table 2**. HF etiology was non-ischemic in 21 subjects (64%) and ischemic in 12 subjects (36%). The median BNP was 454 (IQR 261–1400). On average, transthoracic echocardiogram was conducted 54 days (IQR 16–133) before transplant and hemodynamics were obtained 20 days (IQR 6–40) before transplant. Median LVEF was 19% (IQR 15–27), pulmonary artery pressure was 27 mmHg (IQR 20-35), and pulmonary capillary wedge pressure was 17 mmHg (IQR 11–23). Nearly all patients were receiving inotropes prior to transplant (n=32 [97%]) and a minority were supported with intra-aortic balloon pump (n=7 [21%]). Eight subjects (24%) had RV failure based on PAPi < 1.85. The median PAPi of subjects with RV failure was 1.37 (IQR 0.68 – 1.69) compared to 4.50 (IQR 3.40 – 6.00) among subjects without RV failure. Subjects with RV failure also had a higher median RA/PCWP ratio 0.78 (IQR 0.62 – 0.85) vs 0.34 (IQR 0.22 – 0.45) compared to subjects without RV dysfunction by PAPi (p=0.0003).

**Table 2:** Baseline Clinical Characteristics of the Patients with End-Stage HF.

|  |  |
| --- | --- |
| **Characteristic** | **Overall Cohort**<br>**(n = 33)** |
| Age | 52.7 (61.0 – 44.0) |
| Male Gender | 21 (64%) |
| BMI | 26.5 (24.3 – 30.4) |
| Race/Ethnicity |  |
| White | 26 (79%) |
| Black | 7 (21%) |
| Etiology |  |
| Ischemic | 12 (36%) |
| Non-Ischemic | 21 (64%) |
| Transplant Listing Status |  |
| Status 1A | 10 (30%) |
| Status 1B | 21 (64%) |
| Status 2 | 2 (6%) |
| Pre-Transplant Support |  |
| Milrinone | 32 (97%) |
| IABP | 7 (21%) |
| Laboratory |  |
| BNP (pg/mL) | 454 (261 – 1400) |
| Creatinine (mg/dL) | 1.3 (1.0 – 1.5) |
| Sodium (mEq/L) | 138 (136 – 138) |
| Echocardiographic |  |
| Days before Sample | 54 (16 – 133) |
| LVIDd (cm) | 6.1 (5.4 – 6.9) |
| IVSD (cm) | 0.9 (0.8 – 1.0) |
| LVPWD (cm) | 1.0 (0.8 – 1.1) |
| LVEF | 19 (15 – 27) |
| RV Dilation (Qualitative) |  |
| Normal | 24 (73%) |
| Mild | 6 (18%) |
| Moderate + | 3 (9%) |
| RV Systolic Function (Qualitative) |  |
| Normal | 17 (52%) |
| Mild | 8 (24%) |
| Moderate + | 8 (24%) |
| Hemodynamics |  |
| Days before Sample | 20 (6 – 40) |
| RA mean (mmHg) | 6 (3 – 13) |
| PA Systolic (mmHg) | 39 (31 – 50) |
| PA Diastolic (mmHg) | 18 (12 – 23) |
| PA mean (mmHg) | 27 (20 – 35) |
| PCWP (mmHg) | 17 (11 – 23) |
| Cardiac Output (L/min) | 4.4 (3.8 – 5.1) |
| Cardiac Index (L/min/m<sup>2</sup>) | 2.3 (2.1 – 2.8) |
| PVR (Woods Units) | 2.5 (2.0 – 3.1) |
| PAPi | 3.8 (2.1 – 6.0) |
| BNP: B-type Natriuretic Peptide. IVSd: Interventricular Septal Wall in diastole. LVPWd: Left Ventricular Posterior Wall in diastole. LVIDd: Left Ventricular Internal Diameter in diastole. LVEF: Left Ventricular Ejection Fraction. TAPSE: Tricuspid Annular Plane Systolic Excursion. SBP: Systolic Blood Pressure. DBP: Diastolic Blood Pressure. MAP: Median Arterial Pressure. RA: Right atrium. RV: Right Ventricle. PA: Pulmonary Artery. PCWP: Pulmonary Capillary Wedge Pressure. SVR: Systemic Vascular Resistance. PVR: Pulmonary Vascular Resistance |  |

### Transcriptomic Features of the Right Ventricle under stress in End-Stage Heart Failure

Principal component analysis revealed clear separation between the RV under stress (Compensated RV and Failing RV) compared to Control RV samples, but poorly differentiated between Compensated RV and Failing RV (**Figure 2a**). PLS-DA further supported a distinct separation between the RV under stress and control samples with an improved differentiation between Failing and Compensated RV samples (**Figure 2b**).

**Figure 2A-G:**
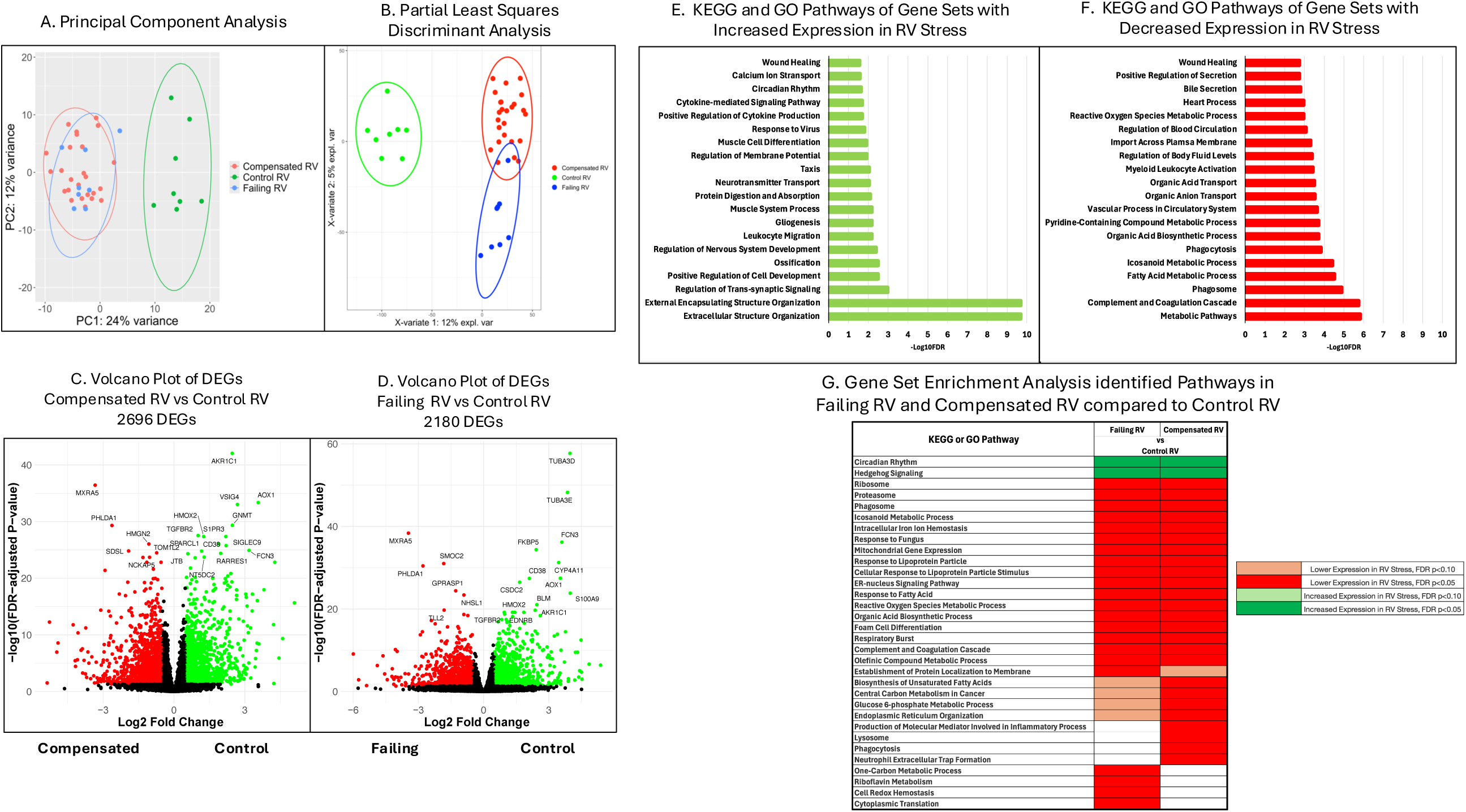
Differential Gene Expression and Pathway Enrichment in the RV under Stress compared to Controls **Figure 2A** displays separation between failing RV, compensated RV, and control RV samples by the first two principal components. On principal component analysis (PCA) there is clear separation between compensated RV samples (red) and control RV samples (green) as well as between failing RV samples (blue) and control samples, however there is poor separation between compensated and failing RV samples. **Figure 2B** displays Partial Least Squares Discriminant Analysis (PLS-DA) based separation between RV sample groups. There is clear separation between control RV samples (green) and compensated RV samples (blue) as well as between control RV samples and failing RV samples (red). PLS-DA based separation between compensated and failing RV samples is significantly improved from PCA. **Figure 2C** displays a Volcano plot for the differentially expressed genes (DEG) comparing between control and compensated RV samples. Genes in green are more highly expressed in control RV samples, while genes in red are more highly expressed in compensated RV samples. **Figure 2D** displays a Volcano plot for the DEGs comparing between control and failing RV samples. Genes in green are more highly expressed in control RV samples, while genes in red are more highly expressed in failing RV samples. **Figure 2E and F** display the top 20 KEGG and GO Pathways enriched among genes with increased expression in RV stress (E) and decreased expression in RV stress (F). **Figure 2G** displays the top 20 KEGG and GO pathways identified on gene set enrichment analysis from the comparisons between failing RV vs control RV (first column) and compensated RV vs control RV (second column). The pathways with higher expression in RV stress are displayed in green, while the pathways with lower expression in RV stress are displayed in red.

We identified 2696 DEGs (1362 with increased counts and 1334 with decreased counts) comparing between Compensated RV vs Control RV and 2180 DEGs (1111 with increased counts and 1069 with decreased counts) comparing between Failing RV vs Control RV (**Figure 2c and 2d**). There were 870 genes with increased counts shared in both comparisons, representing a gene set with increased transcription in the RV under stress. The top 10 genes with increased counts in the RV under stress were MXRA5, PHLDA1, SMOC2, GPRASP2, SDSL, NHSL1, GPRASP1, PHLDB2, KIAA0141, and XPO4. On ORA, the full set of overlapping genes most prominently mapped to the pathways of Extracellular Structure Organization and External Encapsulating Structure Organization (**Figure 2e**). There were 863 genes with decreased counts shared in both comparisons, representing a gene set with decreased transcription in the RV under stress. The top 10 genes with decreased counts in the RV under stress were AOX1, AKR1C1, FCN3, CD38, HMOX2, TGFBR2, GNMT, MARK3, NT5DC2, and JTB. On ORA, the full gene set most prominently mapped to Metabolic Pathways, including Fatty Acid Metabolic Process, Organic Acid Biosynthetic Process, and Icosanoid Metabolic Processes among others (**Figure 2f**). Of note, pathways related to immune system function were identified as both upregulated and downregulated in RV stress.

The top 20 pathways identified on GSEA for comparisons between Failing RV vs Control RV and Compensated RV vs Control RV are shown in **Figure 2g**. Notable trends of differential pathway expression in the RV under stress included under-enrichment of pathways related to protein processing and production (Ribosome, Proteasome, Endoplasmic Reticulum Organization), fatty acid metabolism (Icosanoid Metabolic Process, Response to Lipoprotein Particle, Cellular Response to Lipoprotein Particle Stimulus, Response to Fatty Acid), mitochondrial function (Mitochondrial Gene Expression, Reactive Oxygen Species Metabolic Process, Respiratory Burst) and macrophage function (Phagosome, Foam Cell Differentiation), with over-representation of pathways related to hedgehog signaling and circadian rhythm.

### Transcriptomic Features of Right Ventricular Progression from Compensation to Failure

We then compared transcript counts between the failing RV and compensated RV groups defined by PAPi cutoff of 1.85. The top 10 DEGs were CXCL11, CD209, LOX, SLC25A23, WDR49, EFHC2, COL6A6, LDB3, NFIC, and CA3. Of these only CXCL11, a chemokine, reached our multiple comparison testing and log-fold change significance threshold. We then performed GSEA to identify differentially enriched pathways with the top 20 pathways displayed in **Figure 3**. RV compensation associated most prominently with enrichment in the Ribosome, Proteosome, Cytoplasmic Translation, and Cellular Component Assembly Involved in Morphogenesis pathways. RV failure strongly associated with enrichment of pathways related to inflammation including Response to Protozoan, Complement and Coagulation Cascades, IL-2 Production, and Response to Type II Interferon, among others.

**Figure 3:**
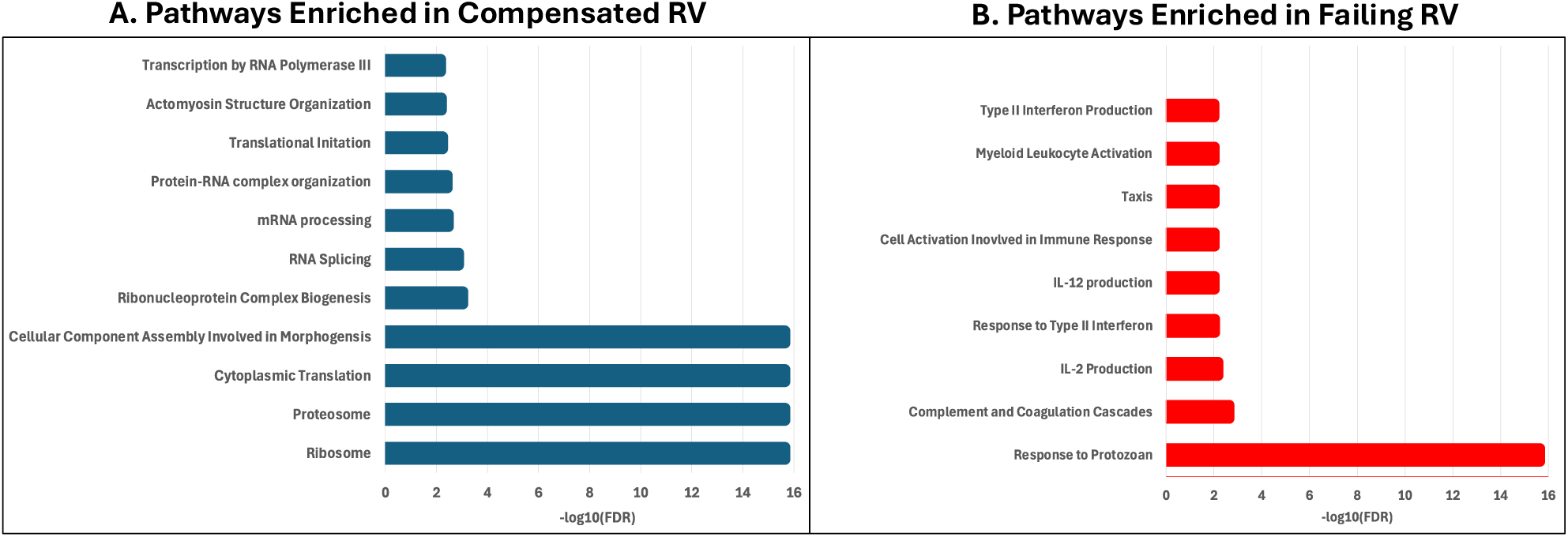
Pathways Enriched in Compensated RV compared to Failing RV identified by Gene Set Enrichment Analysis **Figure 3** displays the top 20 pathways identified on gene set enrichment analysis when comparing compensated RV samples to failing RV samples. **Figure 3A** displays pathways that were found to be enriched in the Compensated RV. **Figure 3B** displays the pathways that were found to be enriched in the Failing RV.

We evaluated whether previously suggested biomarkers of RV failure effectively differentiated between RV control, compensated, and failing tissue in our cohort. We assessed difference in average normalized counts for the extracellular matrix and cell adhesion genes CRTAC1, MEGF9, C1QTNF1, ITGAM, NID1, SPARCL1, TGFBR3, and FAP identified by Khassafi et al^14^ and eight additional proposed biomarkers of RV failure identified by Di Salvo et al^16^ including SERPINA3, SERPINA5, LCN6, LCN10, STEAP4, ARK1C1, STAC2, VSIG4. We found that none of the genes effectively differentiated between failing and compensated RV, however all except CRTAC1 and MEGF9 did effectively differentiate between RV under stress and RV controls (**Figure 4**).

**Figure 4:**
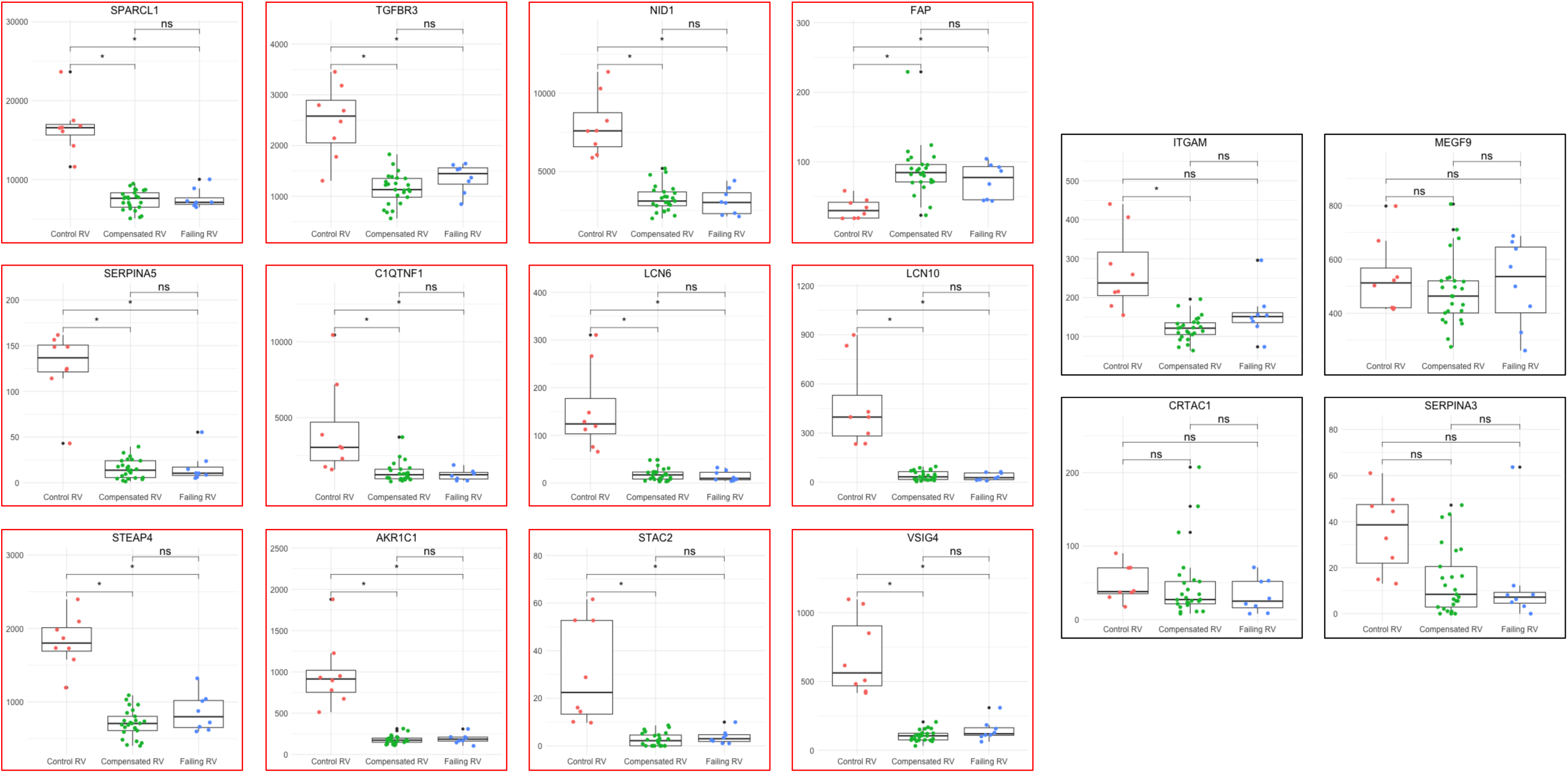
Previously Proposed Biomarker Discrimination between Control, Compensated, and Failing RV samples. **Figure 4** displays boxplots comparing the normalized counts for proposed biomarker genes derived from prior transcriptomic studies comparing between control, compensated, and failing RV samples. We compared normalized counts between groups in a pair-wise fashion by Wilcoxon-Rank Sum Test with Bonferroni correction for multiple comparisons using the number of biomarkers assessed (p-value cutoff of 0.0031 to determine significance). Genes that effectively differentiate between failing RV and control RV as well as compensated RV and control RV are outlined in red. Notably, none of the gene biomarker candidates effectively differentiate between failing and compensated RV samples. ns = non-significant; * = significant.

### Transcriptomic Features of Ventricular Failure Compared Between the Right Ventricle and Left Ventricle

We next sought to understand which transcriptional changes were unique to RV failure in comparison to LV failure. We assessed the overlapping DEGs and GSEA-identified pathways in RV failure (Failing vs Control RV) and LV Failure (Failing vs Control LV). There were 1785 DEGs (791 with decreased counts in failure and 994 with increased counts in failure) comparing between the failing and control LV in patients who had concurrent RV failure (**Figure 5a**). Of these, there were 487 overlapping genes with decreased counts in both RV and LV failure. These genes mapped most prominently to the Metabolic Pathways, Complement and Coagulation Cascades, One-Carbon Metabolic Process, Vascular Process in Circulatory System, and HIF-1 Signaling Pathway gene sets (**Figure 5b**). There were 561 overlapping genes with increased counts in both RV and LV failure. These genes mapped most prominently to Extracellular Structure Organization and External Encapsulating Structure Organization (**Figure 5c**). We then compared between GSEA identified pathways from the failing vs control RV and the failing vs control LV (**Figure 5d**). Control samples in both ventricles were enriched in pathways related to protein transcription and translation (Ribosome, Proteasome, Cytoplasmic Translation, Protein Processing in Endoplasmic Reticulum) as well as mitochondrial function (Cell Redox Hemostasis, Mitochondrial Gene Expression) with increased Hedgehog Signaling. Unique to failure in the RV, there was enrichment of pathways related to fatty acid metabolism and cellular response to lipoprotein/fatty acid (Icosanoid Metabolic Process, Response to Lipoprotein Particle, Cellular Response to Lipoprotein Particle Stimulus, Response to Fatty Acid) in control samples. GSEA-identified pathways unique to LV failure included enrichment of cellular metabolism related gene sets, including Mitochondrial Function (Oxidative Phosphorylation, Mitochondrial Transport, Protein Localization to Mitochondrion, Electron Transport Chain), Glycolysis/Gluconeogenesis, Insulin Signaling Pathway, and protein transcription and translation (Translation Initiation, Protein Neddylation) in control samples. Many of the overarching processes were also implicated in RV failure through different KEGG/GO pathways (i.e Mitochondrial Function, and protein transcription and translation).

**Figure 5:**
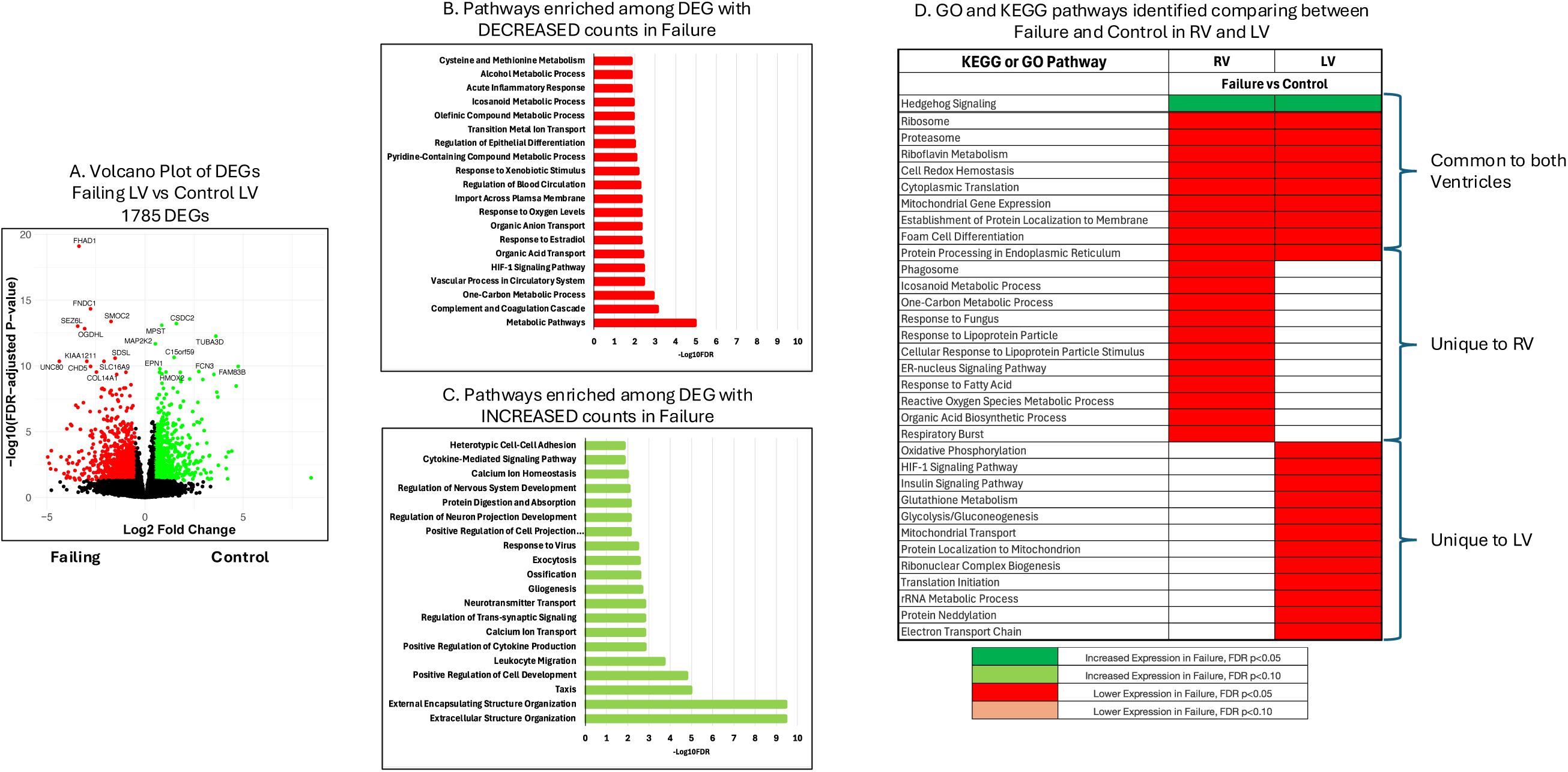
Differential Gene Expression and Pathway Enrichment in Ventricular Failure, Comparing between the RV and LV **Figure 5A** displays a Volcano plot for the differentially expressed genes comparing between Control and Failing LV samples. Genes in green are more highly expressed in control LV samples, while genes in red are more highly expressed in failing RV samples. **Figure 5B** displays the enriched KEGG and GO Pathways among the 487 genes with decreased expression in both RV and LV failure compared to respective controls. **Figure 5C** displays the top 20 enriched KEGG and GO Pathways among the 561 genes with increased expression in both RV and LV failure compared to respective controls. **Figure 5D** displays the top 20 gene set enrichment analysis identified pathways from the comparisons between failing RV vs control RV (first column) and failing LV vs control LV (second column). Pathways with increased expression in failure are displayed in green, while pathways with decreased expression in failure are displayed in red.

### Transcriptomic Differences between the Right and Left Ventricles

We sought to understand which genes and pathways differentiated the RV from the LV in control subjects and HF subjects with biventricular failure. We assessed the overlapping DEGs and GSEA-identified pathways to understand whether differences in RV and LV transcription were similar in control and failing ventricles. PCA demonstrated good separation between failing and control ventricular samples but did not effectively differentiate between RV and LV (**Figure 6a**). Discrimination improved using PLS-DA (**Figure 6b**). We identified 357 DEGs comparing Control RV to Control LV (**Figure 6c**) and 109 DEGs comparing Failing RV to Failing LV (**Figure 6d**). Of these, 36 genes overlapped in both comparisons, representing a gene set that differentiates RV from LV in both control and failure. These genes were most prominently found in metabolism-related pathways including PPAR signaling (ADIPOQ, ANGPTL4, PCK1; p_adj_ = 0.119), Adipocytokine signaling (ADIPOC, PCK1, POMC; p_adj_ = 0.119), carbohydrate biosynthetic process (ADIPOQ, PCK1, GPD1, RBP4; p_adj_ = 0.150), and lipid localization (ADIPOQ, C3, POMC, PRELID2, RBP4; p_adj_= 0.256). By average rank, the top 10 genes differentiating RV and LV regardless of control or failure tissue were CFD, FAIM2, GRXCR2, IGSF10, RBP4, LYVE1, SNAP91, COL28A1, SLC19A3, and DPP6. GSEA comparing RV to LV in both control and failing ventricular myocardium revealed two overlapping pathways, indicating that PPAR signaling and Fatty Acid Metabolism were enriched in RV samples compared to LV samples regardless of control or failing status (Fig 2e). Unique to the failing RV compared to the failing LV, there was an enrichment of pathways related to protein translation and processing (Ribosome, Proteasome, Protein Processing in Endoplasmic Reticulum, Protein Export, Cytoplasmic Translation, Translational Initiation, Protein Neddylation) and mitochondrial function in the RV (Oxidative Phosphorylation, Pyruvate Metabolism, Electron Transport Chain, Mitochondrial Gene Expression, Energy Derivation by Oxidation of Organic Compounds, NADH Dehydrogenase Complex Assembly). In contrast, unique to the control RV compared to the control LV there was a prominent enrichment of inflammation related gene sets.

**Figure 6:**
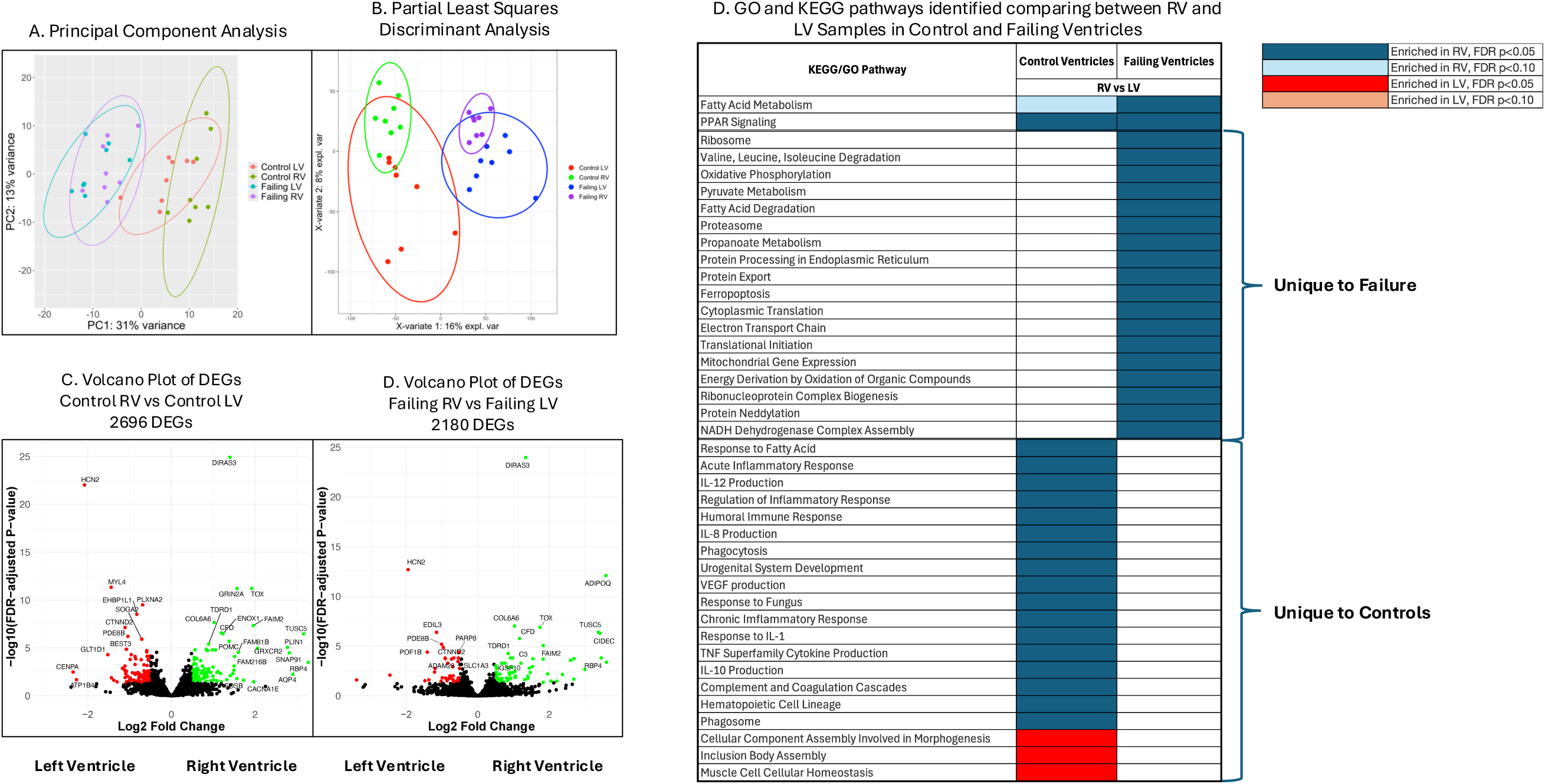
Differential Gene Expression and Pathway Enrichment in The Right Ventricle (RV) Compared to the Left Ventricle (LV) **Figure 6A** displays separation between control RV (Green), failing RV (Purple), control LV (Red), and failing LV (Blue) samples by the first two principal components. There is clear separation between Control and Failing samples, but separation is less clear between RV and LV samples. Figure 6B displays Partial Least Squares Discriminant Analysis (PLS-DA) between control RV (Green), failing RV (Purple), control LV (Red), and failing LV samples (Blue). Separation between control and failing ventricular samples remains robust, with improved separation between RV and LV samples. **Figure 6C and D** display a Volcano plot for the differentially expressed genes (DEG) comparing between the RV and LV in controls (C) and in failure (D). Genes in green are more highly expressed in the RV, while genes in red are more highly expressed in LV samples. **Figure 6E** displays the top 20 KEGG and GO pathways identified on gene set enrichment analysis from the comparisons between RV vs LV in control samples (first column) and failing samples (second column). The pathways with higher expression in the RV are displayed in blue, while the pathways with higher expression in the LV are displayed in red.

Lastly, we assessed whether previously suggested biomarkers of RV failure from Khassafi et al^14^ and Di Salvo et al^16^ were unique to differentiating between RV failure and RV control, or also varied between LV failure and LV control. Of the 16 genes, SPARCL1, TGFBR3, FAP, SERPINA5 and ARK1C1 were differentially expressed between the failing RV and control RV, but not the failing LV and control LV (**Figure 7**). In our study, the top 10 genes that were differentially expressed in the failing RV compared to control RV without differential expression in the failing LV compared to control LV were WASF3, SHB, PATZ1, TRPC6, SPTSSA, IFT43, ART4, LGI3, CTF1, and CPT2 (**Figure 8**). There were no clear KEGG or GO pathways that included multiple genes. Some of these genes could be broadly characterized as associated with lipid metabolism (AKR1C1, CPT2, SPTSSA), cell-cell signaling (TGFBR3, CTF1, LGI3, SHB, ART4, and IFT43), immune function (WASF3, TGFBR3, ART4, SERPINA5), and extracellular matrix remodeling (SPARCL1, TGFBR3, FAP, SERPINA5)

**Figure 7:**
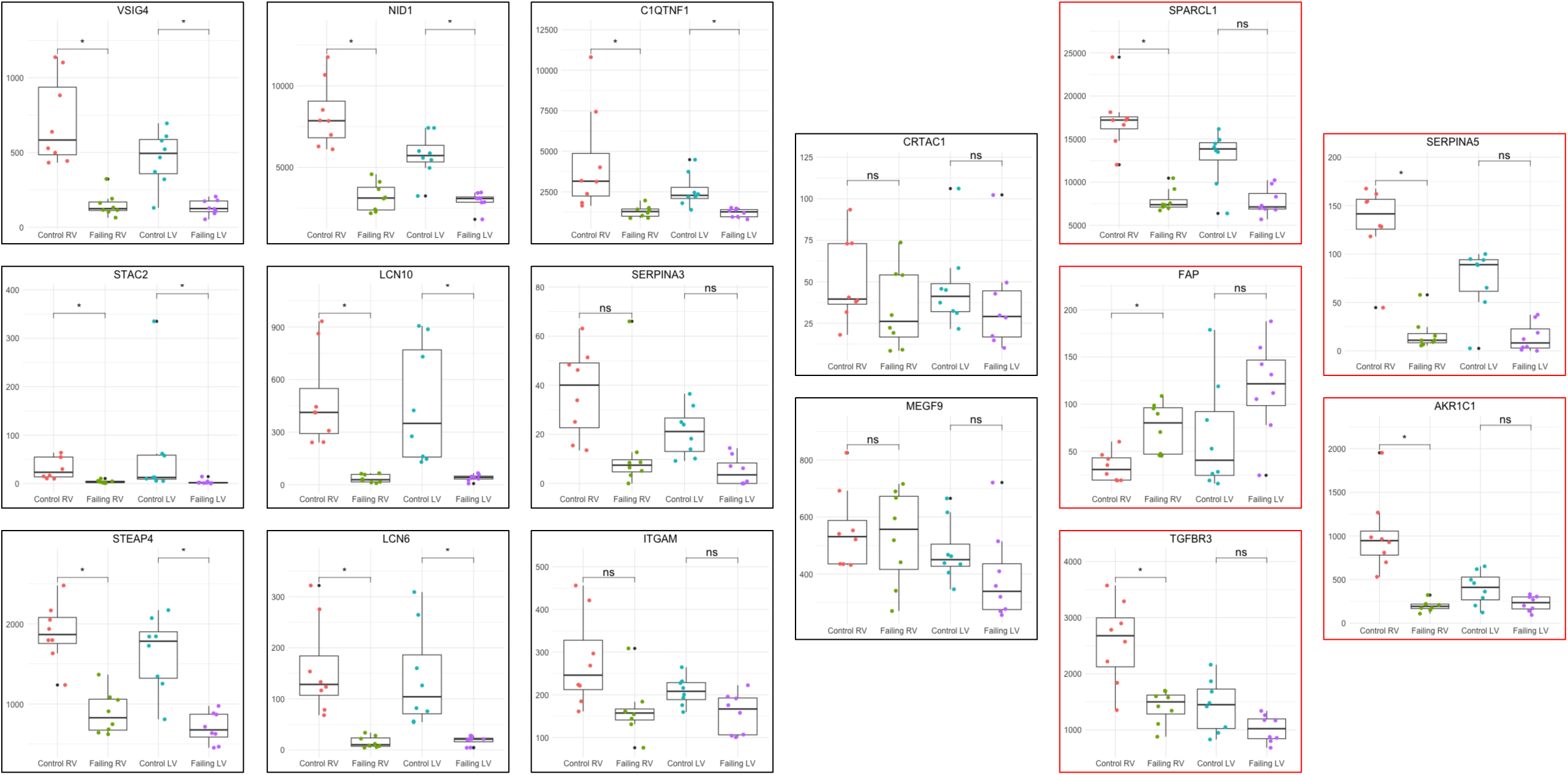
Previously Proposed Biomarker Discrimination of RV Failure compared to LV Failure **Figure 7** displays boxplots comparing the normalized counts for proposed biomarker genes derived from prior transcriptomic studies in control RV, failing RV, control LV, and failing LV. We compared normalized counts between control and failing ventricular samples in the RV and the LV using Wilcoxon-Rank Sum Test with Bonferroni correction for multiple comparisons using the number of biomarkers assessed (p-value cutoff of 0.0031 to determine significance). Genes that effectively differentiate between failing RV and control RV but do NOT change between the failing LV and control LV are outlined in red (SPARCL1, FAP, TGFBR3, SERPINA5, and AKR1C1). ns = non-significant; * = significant.

**Figure 8:**
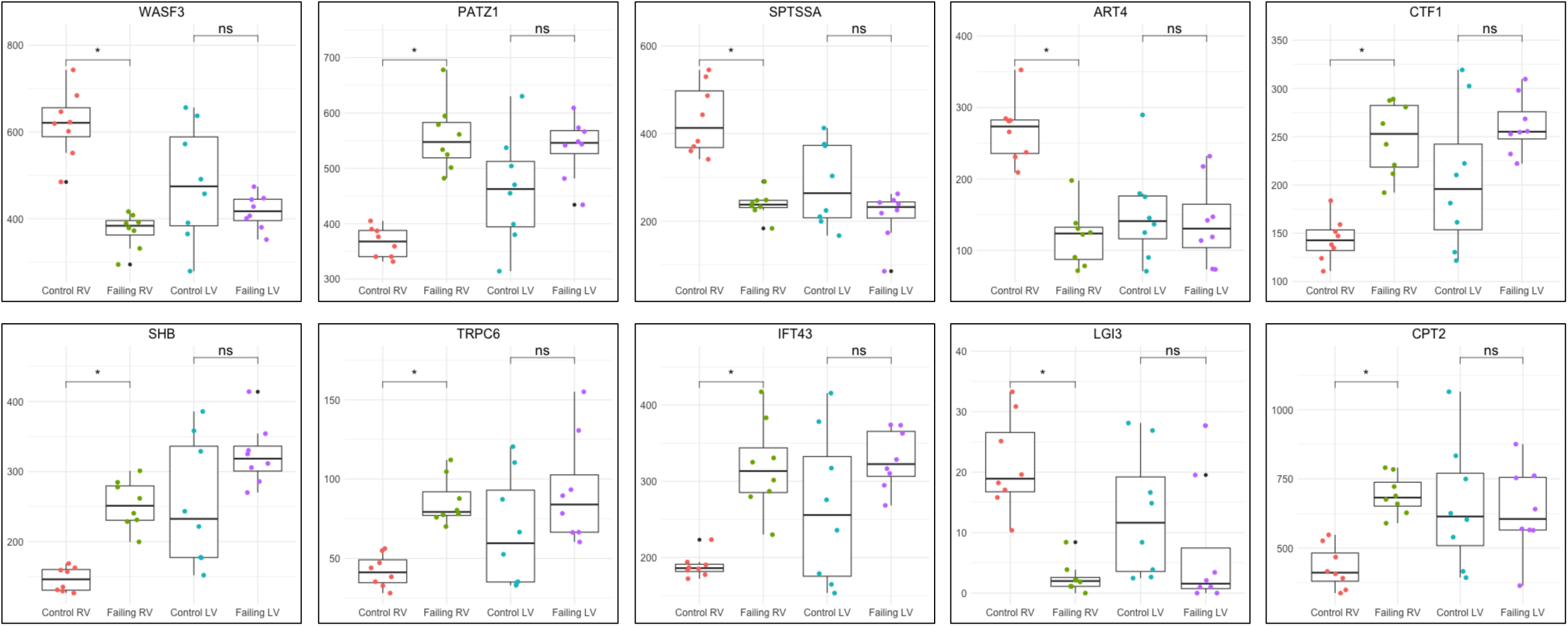
Novel RV-specific Biomarker Candidates **Figure 8** displays boxplots comparing the normalized counts for novel proposed RV-specific biomarker genes. We compared normalized counts between control and failing ventricular samples in the RV and the LV using Wilcoxon-Rank Sum Test with Bonferroni correction for multiple comparisons using the number of biomarkers assessed (p-value cutoff of 0.005 to determine significance). ns = non-significant; * = significant.

## Discussion

In this study, we performed bulk myocardial RNA-sequencing of paired LV and RV samples from patients with end-stage HF and controls. We then employed transcriptomic analyses including differential gene expression and enrichment analysis to identify transcriptional changes in the RV under stress and to distinguish between compensation and failure. We compared these findings to the transcriptional changes in LV failure and assessed differences in the transcriptional signatures of the RV and LV. The major findings are: 1) The RV myocardium under the stress of LV failure exhibits transcriptional changes highly similar to those identified in pre-capillary PH related RV failure, 2) Gene sets related to inflammation and protein production/processing differentiate between compensated RV and failing RV, 3) PPAR signaling and fatty acid metabolism gene sets are consistently enriched in the RV compared to the LV regardless of control or failure status. Moreover, fatty acid metabolism is implicated as the primary differentiator between transcriptomic signatures of RV failure and LV failure, 4) The transcriptional profile of the control RV features prominent enrichment of inflammation and immune system pathways compared to the LV, 5) We identify potential transcriptomic biomarkers of RV failure that are specific to the RV. These findings identify unique biology and pathology in the human RV and may contribute to development of RV-specific biomarkers and therapeutics in HF, the most common cause of RV dysfunction and failure.

The transcriptional changes we found in the RV tissue of end-stage HF subjects compared to controls suggest that similar molecular mechanisms may underly RV dysfunction in both pre-capillary PH and end-stage HF. Comparable myocardial transcriptomic studies in human PAH and multiple animal models that recapitulate PAH via pre-capillary pulmonary hypertension (Pulmonary artery banding, Sugen/Hypoxia, and Monocrotaline) have consistently identified an association between RV failure and increased expression of genes related to ECM remodeling, decreased expression of genes related to fatty acid metabolism and mitochondrial function, and variable change in expression of genes related to inflammation.^14,26–28^ Furthermore, targeted investigations have validated that these transcriptional changes correspond to increased ECM protein expression,^14,29^ impaired fatty acid oxidation,^13,30^ and dysregulated inflammation^7^ in human subjects along with decreased mitochondrial function in rats.^31^ Although transcriptomic findings must be validated as translating to protein expression, our findings suggest that molecular pathways identified in pre-capillary PH may also be areas for the development of biomarkers and therapeutics in HF-associated RV dysfunction.

Our study is unique from many prior transcriptomic investigations of RV dysfunction in attempting to discriminate between the compensated and failing RV, with several strengths over the few prior studies examining this question. Two studies have examined RV transcriptomic changes comparing between compensated and failing RV in patients with end-stage LV failure, finding few DEGs.^16,32^ The first was conducted in 22 subjects with end-stage HF and defined RV failure solely by echocardiographic variables with few patients meeting hemodynamic criteria and substantial time between echocardiogram and explant (mean 98 +/-85 days).^16^ The second included only 6 subjects with an LV assist device in place and employed solely hemodynamic criteria (elevated right sided filling pressure [>18 mmHg] and low cardiac index [<2.0 L/min/m^2^]), further categorizing severity by need for inotropic or mechanical support.^32^ One other study performed RNA-seq on human compensated RV samples from 15 subjects with congenital heart disease obtained at biopsy, confirmed to have normal RV function by cardiac index (>2.2 L/min/m^2^) and tricuspid annular plane systolic excursion (17 mm), comparing these to failing RV samples from 11 subjects with PAH obtained at autopsy.^14^ They identified 260 DEGs that implicated enrichment of mitochondrial function associated pathways, however these results may be biased by inherent differences in myocardial transcription between congenital heart disease and PAH. In contrast, our study was the largest to date with 33 subjects and employed a robust clinically and pathologically validated hemodynamic definition of RV failure that was obtained shortly prior to explant. Similar to the studies conducted in end-stage HF, we found few DEGs but present new GSEA findings that implicate protein transcription and translation pathway enrichment in RV compensation and cytokine production/signaling as well as leukocyte activation pathway enrichment in RV failure. The limited differential gene expression noted in our study and prior investigations may relate to the criteria employed to differentiate between compensated and failing RV, limited sample sizes, or the heterogeneity of subjects.

The second unique aspect of our study was the direct comparison of transcriptional changes in RV failure and LV failure using paired RV and LV samples from control subjects and subjects with objectively defined biventricular failure. Although prior studies have separately examined transcriptional changes in each RV failure and LV failure, the findings from separate studies are challenging to consolidate due to differing definitions of RV failure, clinically heterogeneous populations, and differences in analytic strategies. Given the differences in embryological origin and physiological pump conditions of the right and left ventricle,^33^ we expected to find transcriptional differences. There was a notable enrichment of gene sets related to lipid signaling and metabolism in RV failure that were not noted in the LV failure. In addition, transcriptional differences between the RV and LV in both control and failure comparisons identified Fatty Acid metabolism, PPAR signaling, and Adipocytokine signaling as pathways enriched in the RV compared to LV. These findings suggest constituent differences between RV and LV metabolism involving fatty acids and associated metabolic regulatory signaling with changes to fatty acid metabolism that affect the RV in failure more prominently than the LV. Dysregulation of fatty acid metabolism is strongly implicated in RV failure related to PAH^6,13,30,34^ and in LV failure,^35^ however fatty acid oxidation exhibits variable change in the failing LV, while it is consistently decreased in PAH-associated RV failure. Considered altogether, these findings suggest that certain changes to fatty acid metabolism and related signaling may be unique to the RV and potential areas for RV-specific biomarker development. We found that failure in both ventricles was associated with mitochondrial dysfunction, decreased protein transcription and translation, and changes to cellular metabolism, aligning with prior studies.^14,36^ We also noted enrichment of sonic hedgehog signaling in both RV and LV failure, a pathway that has not been noted in transcriptomic studies but has been implicated in cardiac regeneration and endothelial cell activation following injury.^37–39^

Direct comparison of transcriptional profiles between the RV and LV in controls and failure identified enrichment of inflammation and immune function as the predominant differentiating pathway in the control ventricles and mitochondrial function as the predominant differentiating pathway in the failing ventricles. Inflammation and immune function related pathways were enriched in the control RV compared to LV, which may be due to transcriptional differences, but may be related to differential cell composition as animal studies have demonstrated a higher proportion of macrophages in the RV than the LV.^40^ There is evidence that a dysfunctional immune system with myocardial inflammation plays a role in both ventricles when failing,^41,42^ however, whether these changes are similar in the failing RV and failing LV remains an outstanding question. Mitochondrial function related pathways were enriched in the failing RV compared to the failing LV. Rodent bioenergetic studies suggest increased oxygen consumption due to increased fatty acid oxidation in the normal RV cardiomyocytes compared to LV cardiomyocytes,^43^ however we did not find a transcriptional difference in mitochondrial function related pathways comparing human control RV to control LV. Mitochondrial dysfunction may play a relatively less significant role in RV failure compared to LV failure, however, there are no metrics to directly compare the degrees of dysfunction in the RV and LV and this difference in expression may related to a greater degrees of “failure” in LV samples compared to RV samples.

Development of a RV failure-specific biomarker in subjects with LV failure presents a unique challenge given the overlap in pathophysiology between the ventricles. We found that most previously suggested biomarkers for RV failure demonstrated a parallel change when comparing between failing and control LV samples, suggesting that these biomarkers were not specific to the RV and therefore likely to be affect by functional changes to both ventricles. Our study identified 13 genes that are plausibly associated with RV failure and are differentially expressed in RV failure, but not LV failure. Ultimately, these genes and the pathways implicated may correlate with measurable plasma levels of transcript, protein or metabolite that can specifically identify RV failure in patients with HF from LV failure.

### Limitations

As our study employed bulk tissue RNA-sequencing, we are not able to determine the relative contribution of different cell types or compare transcription between individual cell types in each ventricle, both of which are factors that likely contribute to tissue function. Transcriptomic based analysis leverages relative transcript counts for biological inferences, which ultimately may not translate to protein expression or cellular/tissue function, therefore all findings must be considered within this context. We selected a well-accepted definition of right ventricular failure/dysfunction that is prognostically and pathologically validated (PAPi <1.85) in end-stage heart failure, however there is no singular accepted definition, and it is possible that this definition misclassified samples. Because this study included a limited number of human samples, and we were not able to adjust our analyses for patient characteristics that might introduce bias including age, sex, demographics, or underlying etiology of HF. We included subjects with ischemic and non-ischemic etiologies of HF, which may impact our findings. Nearly all patients with HF in this study were actively receiving chronic inotropic support with milrinone, which may affect myocardial transcription.

## Conclusion

It is likely that the molecular mechanisms of PAH and HF-associated RV dysfunction have significant overlap. Larger studies with rigorous, prospective assessment of RV function are necessary to define the molecular processes associated with the progression from RV compensation to failure. Fatty Acid metabolism, PPAR signaling, and Adiopokine signaling are enriched in the RV compared to the LV and may be promising pathways for RV-specific biomarker development.

## Data Availability

All data produced in the present study are available upon reasonable request to the authors.

## Acknowledgements

The Vanderbilt VANTAGE Core provided technical assistance for this work. VANTAGE is supported in part by CTSA Grant (5UL1 RR024975-03), the Vanderbilt Ingram Cancer Center (P30 CA68485), the Vanderbilt Vision Center (P30 EY08126), and NIH/NCRR (G20 RR030956). Myocardial samples were obtained from the Vanderbilt Cardiology Core Laboratory for Translational and Clinical Research.

